# Recording of “COVID-19 vaccine declined” among vaccination priority groups: a cohort study on 57.9 million NHS patients’ primary care records in situ using OpenSAFELY

**DOI:** 10.1101/2021.08.05.21259863

**Authors:** Helen J Curtis, Peter Inglesby, Brian MacKenna, Richard Croker, William Hulme, Christopher T Rentsch, Krishnan Bhaskaran, Alex J Walker, Caroline E Morton, David Evans, Amir Mehrkar, Seb Bacon, Chris Bates, George Hickman, Tom Ward, Jessica Morley, Jonathan Cockburn, Simon Davy, Anna Schultze, Elizabeth Williamson, Helen I McDonald, Laurie Tomlinson, Rohini Mathur, Rosalind M Eggo, Kevin Wing, Angel YS Wong, Harriet Forbes, John Tazare, John Parry, Frank Hester, Sam Harper, Shaun O’Hanlon, Alex Eavis, Richard Jarvis, Dima Avramov, Paul Griffiths, Aaron Fowles, Nasreen Parkes, Stephen JW Evans, Ian J Douglas, Liam Smeeth, Ben Goldacre

## Abstract

**Background:** All patients in England within vaccine priority groups were offered a COVID-19 vaccine by mid-April 2021. Clinical record systems contain codes to denote when such an offer has been declined by a patient (although these can in some cases be entered for a variety of other reasons including vaccination delay, or other administrative issues). We set out to describe the patterns of usage of codes for COVID-19 vaccines being declined.

**Methods:** With the approval of NHS England and using the full pseudonymised primary care records for 57.9 million NHS patients, we identified all patients in key vaccine priority groups: aged over 50, or over 16 and at increased risk from COVID-19 (Clinically Extremely Vulnerable [CEV] or otherwise “at risk”). We describe the proportion of patients recorded as declining a COVID-19 vaccination for each priority group, and by other clinical and demographic factors; whether patients recorded as “declined” subsequently went on to receive a vaccination; and the distribution of code usage across GP practices.

**Results:** Of 24.5 million patients in priority groups as of May 25th 2021, 89.2% had received a vaccine, 8.8% had neither a vaccination nor a decline recorded, and 663,033 (2.7%) had a decline code recorded. Of patients with a recorded decline, 125,587 (18.9%) were subsequently vaccinated. Subsequent vaccination was slightly more common in the South Asian population than other ethnicities (e.g. 32.3% vs 22.8%, over 65s). The proportion of declining-unvaccinated patients varied strongly with ethnicity (Black 15.3%, South Asian 5.6%, White 1.5% in over 80s); and was higher in patients from more deprived areas. COVID-19 vaccine decline codes were present in almost all practices (98.8%), but with wide variation between practices in rates of usage. Among all priority groups, declining-unvaccinated status was most common in CEV (3.3%).

**Conclusions:** Clinical codes indicative of COVID-19 vaccinations being declined are widely used in English general practice. They are substantially more common among Black and South Asian patients, and patients from more deprived areas. There is a need for more detailed survey and/or qualitative research with patients and clinicians to determine the most common reasons for these recorded declines.

## Background

On December 8th 2020, the NHS in England administered the first COVID-19 vaccination as part of an ambitious vaccine programme to combat the ongoing COVID-19 pandemic. All people in England within the initial Joint Committee on Vaccination and Immunisation (JCVI) vaccine priority groups (box 1), had been invited to get a COVID-19 vaccine by the middle of April (NHS England 2021), after which invitations were extended to all other adults. We have previously described the detailed trends and clinical characteristics of COVID-19 vaccine recipients using 57.9 million patients’ records (The OpenSAFELY Collaborative et al. 2021) and we update a weekly report covering 40% of the population (<u>link</u>). The campaign vaccinated 19 out of every 20 people aged over 50, but concerns remain around lower vaccine coverage in some groups, particularly ethnic minorities (NHS-England 2021; The OpenSAFELY Collaborative et al. 2021).

Electronic health record (EHR) software has functionality to record when a vaccination has been declined, and SNOMED-CT, the mandated coding language in NHS primary care, has several codes that may be used for this purpose (Table S2a). These codes may be used where a patient has explicitly and absolutely refused a vaccine; however they may also sometimes be used for other reasons, such as when a patient wishes to delay getting the vaccine (e.g. due to illness), or rejects a vaccine invitation from one organisation after booking an appointment to be vaccinated elsewhere. An additional range of codes are available to indicate other situations including contraindications, vaccination appointments being missed, or the vaccine being “not given” (Table S2b), but their usage may occasionally cross over. As there is no comprehensive national guidance or specification on how practices should use these codes, individual practices may also use them to facilitate the organisational delivery of this large-scale vaccination campaign. For example, in order to prevent further automated invites, a practice may add a code indicating a patient has declined when no response has been received after a certain number of invitations, or they may be used in uncertain circumstances such as a possible intolerance.

We therefore set out to describe the patterns in recorded COVID-19 vaccine declines among 57.9 million pseudonymised patient records (∼95% of registered patients in England) held in the OpenSAFELY platform, a new secure analytics platform for NHS patient data.

## Methods

### Study design

We conducted a retrospective cohort study using general practice primary care electronic health record (EHR) data from all England general practices supplied by the EHR vendors EMIS and TPP. Follow-up began on 8th December 2020, the start of the national vaccination campaign, and ended on May 25th 2021, the latest available at the time of analysis and more than one month after all those in priority groups had been offered a vaccination (NHS England 2021).

### Study population

We included all patients registered with a general practice in England using EMIS or TPP software on May 25th 2021 and identified as belonging to a vaccine priority group (box 1). We additionally excluded patients with unknown date of birth (i.e. age >120), or with unknown sex.

### Data Source

Primary care records managed by the GP software providers EMIS and TPP were accessed through OpenSAFELY, an open source data analytics platform created by our team on behalf of NHS England to address urgent COVID-19 research questions (https://opensafely.org). OpenSAFELY provides a secure software interface allowing a federated analysis of pseudonymized primary care patient records from England in near real-time within the EMIS and TPP highly secure data environments. Non-disclosive, aggregated results are exported to GitHub where further data processing and analysis takes place. This avoids the need for large volumes of potentially disclosive pseudonymised patient data to be transferred off-site. This, in addition to other technical and organisational controls, minimizes any risk of re-identification. The dataset available to the platform includes pseudonymised data such as coded diagnoses, medications and physiological parameters. No free text data are included. All activity on the platform is publicly logged and all analytic code and supporting clinical coding lists are automatically published. In addition, the framework provides assurance that the analysis is reproducible and reusable. Further details on our information governance and platform can be found below under <u>information governance and ethics</u>.

### COVID-19 vaccine status

Vaccine administration details are recorded in the National Immunisation Management Service (NIMS) and electronically transmitted to every individual’s GP record on a daily basis. We ascertained which patients had any recorded COVID-19 vaccine administration code in their primary care record. We also captured other clinical codes for a COVID-19 vaccination which may have been entered outside of the usual system (Table S1). Patients were considered to be vaccinated if any COVID-19 vaccination record or code was present, irrespective of the number of doses received.

### Vaccines declines and other reasons for not being vaccinated

In March 2021, NHS Digital published a news article listing COVID-19 vaccination codes (NHS Digital 2021). From this document we identified all codes indicative of declining a COVID-19 vaccine as those containing the word “Declined” in the description (Table S2a). Additionally we included three further codes fitting this pattern, either reported in the national COVID-19 Vaccination Uptake Reporting Specification (“COVID-19 Vaccination Uptake Reporting Specification” n.d.), or inactive codes. Patients were assigned to the Declined group if they had any code for a COVID-19 vaccination being Declined, irrespective of their vaccination status. We describe subsets of the Declined group as: those who had already had a vaccination (Declined Post-vaccination); those who later received their first dose (Declined Then Received); and those with no recorded vaccination (Remaining Declined). In patients with no recorded vaccination or Declined code we looked for any other records indicating an attempt or intention to be vaccinated, e.g. a contraindication or “did not attend”, Table S2b) and assigned these patients to the “Contraindicated/Unsuccessful” group. All other unvaccinated patients were assigned to the “No Records” group.

### Priority groups for vaccination

We classified patients into priority groups (box 1) using SNOMED-CT codelists and logic defined in the national *COVID-19 Vaccination Uptake Reporting Specification* developed by PRIMIS v1.1 (“COVID-19 Vaccination Uptake Reporting Specification” n.d.). However, in order to report age groups and clinical groups separately, here we combine the 70-74 and 75-79 cohorts together as group 3, leaving the CEV cohort separate as group 4; we also limited the care home population to those aged 65+ (box 1). We did not assess eligibility as defined by occupation, i.e. health and care staff for the relevant priority groups (1 and 2) because this information is largely missing or unreliable in GP records. These patients would be classified into a lower priority group where applicable (e.g. by clinical risk or age) and are otherwise excluded. Each patient was assigned only to their highest priority group and not included again as part of any other priority group. In line with the national reporting specification, age was calculated as at 31 March 2021 while other criteria were ascertained using the latest available data at the time of analysis.

#### Box 1

**Priority groups for vaccination advised by the Joint Committee on Vaccination and Immunisation (JCVI)**(Public Health England 2020), and the priority groups used in this report. The final column indicates how priority groups are combined into three larger groups where this was necessary for data presentation. Although pregnant women were not included in the CEV or At Risk groups on the basis of their pregnancy, some pregnant women will be included in these groups based on other criteria.

| Priority Group | JCVI Risk group | Groupings used in this report |  |  |
| --- | --- | --- | --- | --- |
|  |  | Priority group | Group name | Combined group |
| 1 | Residents in a care home for older adults<br>Staff working in care homes for older adults | Residents in a care home for older adults, **aged 65+** | Care home | 65+ |
| 2 | All those 80 years of age and over<br>Frontline Health and social care workers | All those 80 years of age and over | 80+ |  |
| 3 | All those 75 years of age and over | All those **70** years of age and over | 70-79 |  |
| 4 | All those 70 years of age and over |  |  |  |

|  | Clinically extremely vulnerable individuals (not including pregnant women and those under 16 years of age) | Clinically extremely vulnerable individuals (not including pregnant women and those under 16 years of age) | CEV | CEV / At Risk |
| --- | --- | --- | --- | --- |
| 5 | All those 65 years of age and over |  | 65-69 | 65+ |
| 6 | Adults aged 16 to 65 years in an at-risk group |  | At Risk | CEV / At Risk |
| 7 | All those 60 years of age and over |  | 60-64 | 50-64 |
| 8 | All those 55 years of age and over |  | 55-59 |  |
| 9 | All those 50 years of age and over |  | 50-54 |  |

### Key demographic and clinical characteristics

We extracted all patient demographics defined by the national reporting specification (for example, ethnicity). We made a small modification to the pregnancy flag, restricting this to females aged under 50, to avoid including any codes incorrectly recorded against males and post-menopausal women. We also extracted demographics not defined by the specification, including the Index of Multiple Deprivation (IMD; 2019 values), derived from patient postcodes at Lower Super Output Area level, grouped into quintiles.

### Codelists and implementation

Information on all characteristics were obtained from primary care records by searching TPP SystmOne and EMIS records for specific coded data. EMIS and TPP SystmOne are fully compliant with the mandated NHS standard of SNOMED-CT clinical terminology. Medicines are entered or prescribed in a format compliant with the NHS Dictionary of Medicines and Devices (dm+d) (MacKenna 2019). Codelists and logic for most features in the national reporting specification were automatically converted to software (https://codelists.opensafely.org/codelist/primis-covid19-vacc-uptake/).

### Missing data

Patients with missing ethnicity or IMD information are included as “Unknown”. A very small number of patients’ vaccinations (0.0012%) or declines (0.051%) were dated before the start of the vaccination campaign or lacked a date altogether; accuracy was prioritised in determining whether a decline was recorded prior to a vaccination, so these patients where the precise sequence could not be determined were counted in the “declined post-vaccination” group. Codes specifically relating to vaccine allergy or contraindications could not be retrieved from the EMIS system, so a small number of affected patients will be counted in the unvaccinated “No records” group.

### Study Measures

We calculated the daily cumulative number and rate of COVID-19 vaccinations, coded vaccine declines, those with contraindications/unsuccessful vaccinations and those with no records related to vaccination, for each priority group. We also measured how many people were vaccinated after previously being recorded to have declined, and the time between these records (0-<2 weeks, 2-<4 weeks, 1-<2 months, ≥ months). We present time trends, bar charts and brief descriptive statistics. We assess the rate of declines recorded at practice level per thousand patients, excluding practices with 250 or fewer registered patients in priority groups and those with less than 10 vaccinated patients, and present this as a histogram and heatmap. Patient counts were rounded to the nearest seven and values under 7 suppressed prior to release from each EHR system; practice counts of 1-3 were shown as 2.

### Software and Reproducibility

Data management and analysis was performed using the OpenSAFELY software libraries and Python, both implemented using Python 3. This analysis was delivered using federated analysis through the OpenSAFELY platform: codelists and code for data management and data analysis were specified once using the OpenSAFELY tools; then transmitted securely to the OpenSAFELY-TPP platform within TPP’s secure environment, and separately to the OpenSAFELY-EMIS platform within EMIS’s secure environment, where they were each executed separately against local patient data; summary results were then reviewed for disclosiveness, released, and combined for the final outputs. All code for the OpenSAFELY platform for data management, analysis and secure code execution is shared for review and re-use under open licenses at GitHub.com/OpenSAFELY. All code for data management and analysis for this paper is shared for scientific review and re-use under open licenses on GitHub (https://github.com/opensafely/covid-vaccine-not-received).

### Patient and Public Involvement

Patients were not formally involved in developing this specific exploratory study that was produced rapidly in the context of the rapid vaccine rollout during a global health emergency. We have developed a publicly available website https://opensafely.org/ through which we invite any patient or member of the public to contact us regarding this study or the broader OpenSAFELY project.

## Results

Of 57.9 million patients in total, 24.5 million were identified as being in priority groups, of whom 21.8 million (89.2%) had received at least one COVID-19 vaccine by 25th May 2021 (Table 1). Some 663,033 (2.7%) were recorded with a code suggestive of declining a vaccine, 125,587 (18.9%) of whom were later vaccinated, while 8.1% had already had a vaccination (Table 1). Thus, 483,791 (2.0%) of people in priority groups have been recorded as declining and remain unvaccinated. Only 15,015 patients (0.1%) had no vaccine or decline recorded but had a recorded contraindication or unsuccessful vaccination (e.g. did not attend appointment), while a further 8.8% had no records of vaccination, decline, contraindication or other vaccine-related codes.

**Table 1.** Summary of COVID-19 vaccination status and declines recorded for patients in OpenSAFELY by priority group as at 25th May 2021.

| Group | Vaccinated (%) | Declined |  |  |  | Contraindicated/ Unsuccessful (%) | No records (%) | Total |
| --- | --- | --- | --- | --- | --- | --- | --- | --- |
|  |  | Total Declined (%) | Declined Then Received (%) | Declined post-vaccine (%) | Remaining Declined (%) |  |  |  |
| Care home | 96.1 | 2.5 | 0.6 | 0.3 | 1.6 | 0.0 | 2.3 | 252,637 |
| 80+ | 95.7 | 3.4 | 1.0 | 0.2 | 2.1 | 0.0 | 2.2 | 2,578,870 |
| 70-79 | 95.3 | 2.7 | 0.6 | 0.4 | 1.8 | 0.1 | 2.9 | 4,754,792 |
| CEV | 87.1 | 4.4 | 0.8 | 0.3 | 3.3 | 0.1 | 9.6 | 1,991,549 |
| 65-69 | 92.4 | 2.3 | 0.3 | 0.2 | 1.8 | 0.1 | 5.8 | 2,503,298 |
| At Risk | 83.1 | 3.0 | 0.4 | 0.1 | 2.4 | 0.1 | 14.4 | 4,324,663 |
| 60-64 | 88.8 | 2.1 | 0.3 | 0.2 | 1.6 | 0.1 | 9.5 | 2,152,038 |
| 55-59 | 86.5 | 2.2 | 0.4 | 0.2 | 1.5 | 0.1 | 11.9 | 2,804,333 |
| 50-54 | 83.7 | 2.0 | 0.3 | 0.1 | 1.6 | 0.1 | 14.7 | 3,114,629 |
| All | 89.2 | 2.7 | 0.5 | 0.2 | 2.0 | 0.1 | 8.8 | 24,476,809 |

CEV: Clinically Extremely Vulnerable. "Vaccinated" includes patients who were previously recorded to have declined (these are also shown separately in the "Declined then Received" column). "Contraindicated/ Unsuccessful" includes patients with no recorded vaccination or Declined code, but with any other code indicative of contraindication or an unsuccessful attempt to vaccinate, e.g. "did not attend" (Appendix Table S1). Patient counts are rounded to the nearest 7.

### Individual priority groups

The total rate of declines being recorded was highest in the CEV group at 4.4%, followed by 80+ (3.4%), and At Risk (3.0%) (Table 1). Removing those who were vaccinated, CEV remained the highest with 3.3% recorded as declining (−1.1% absolute reduction), with 80+ reducing to 2.1% (−1.3%), with a smaller reduction for the At Risk group to 2.4% (−0.6%) (Figure 1–2, Table S1). Within the CEV/At Risk groups there was a strong correlation with age group, and comparing each five-year age band with other priority groups, the percentage of people recorded as declining was still higher in the CEV/At Risk group, e.g. 1.88% vs 1.56% for ages 60-64 (Table S3); however, the percentage vaccinated in each age band was also higher. Patients recorded as declining a COVID-19 vaccination accounted for approximately half of those currently unvaccinated amongst the 80+ group, and more than one third in the other three top eligibility groups (Care Home, 70-79 and CEV; Figure 1, Table 1).

**Figure 1.**
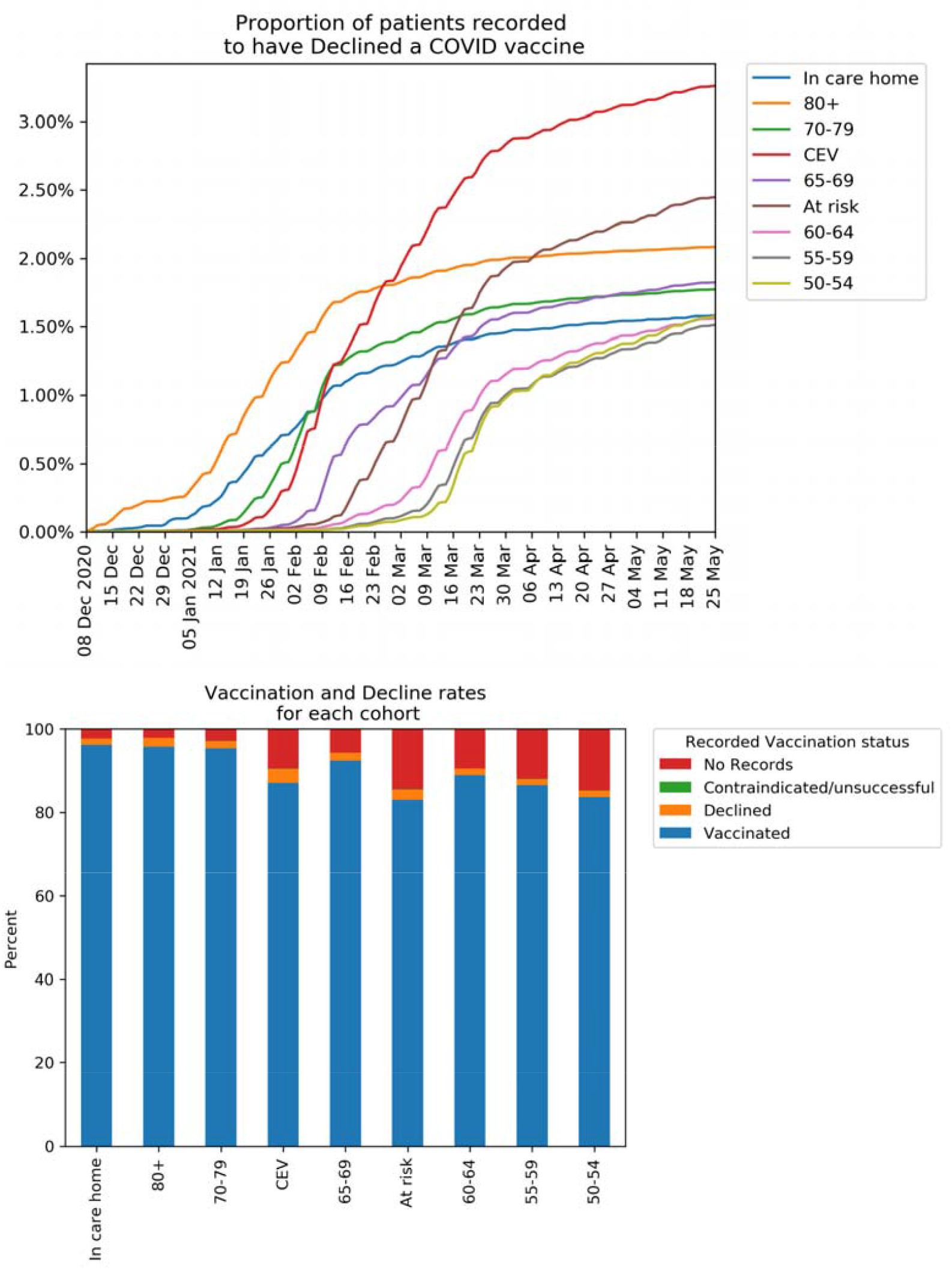
Recorded vaccination status of patients in OpenSAFELY up to May 25th 2021. (a) Cumulative percentage of patients in each priority group recorded as declining a COVID-19 vaccination and remaining unvaccinated. (b) Recorded COVID-19 vaccination status for patients by priority group. “Declined” excludes patients with a recorded vaccination. “Vaccinated” includes those previously recorded as declining.

### Variation by demographic factors

The percentage of those in each ethnic group who had a decline recorded and are unvaccinated, split by individual priority group, is shown in figure 2a, with time trends in figure S1 by combined priority groups (65+, CEV/At Risk, 50-64). The percentage of the White population who were recorded as declining (and unvaccinated) was similar across each priority group (1.3%-1.5%), except for CEV and At Risk groups which were slightly higher at 2.7% and 2.3% respectively (figure 2a); the variation in most other ethnic groups was more marked, especially in the Black population. The highest rate within the Black population was 15.3% (aged 80+), more than ten times greater than the White 80+ population (1.5%), while the lowest rate was 3.2% (aged 50-54), higher than any of the groups in the White population. The percentage in the South Asian population was generally lower than other non-White groups, ranging from 1.4% (aged 50-54) to 5.6% (aged 80+) (figure 2a). Time trends charts show that these differences have been consistent but increasing over time (figure S1a-c). There was also a much larger proportion of people in each ethnic minority group with no records of vaccination with no reason recorded compared to the White population (figure 3, table S4).

**Figure 2.**
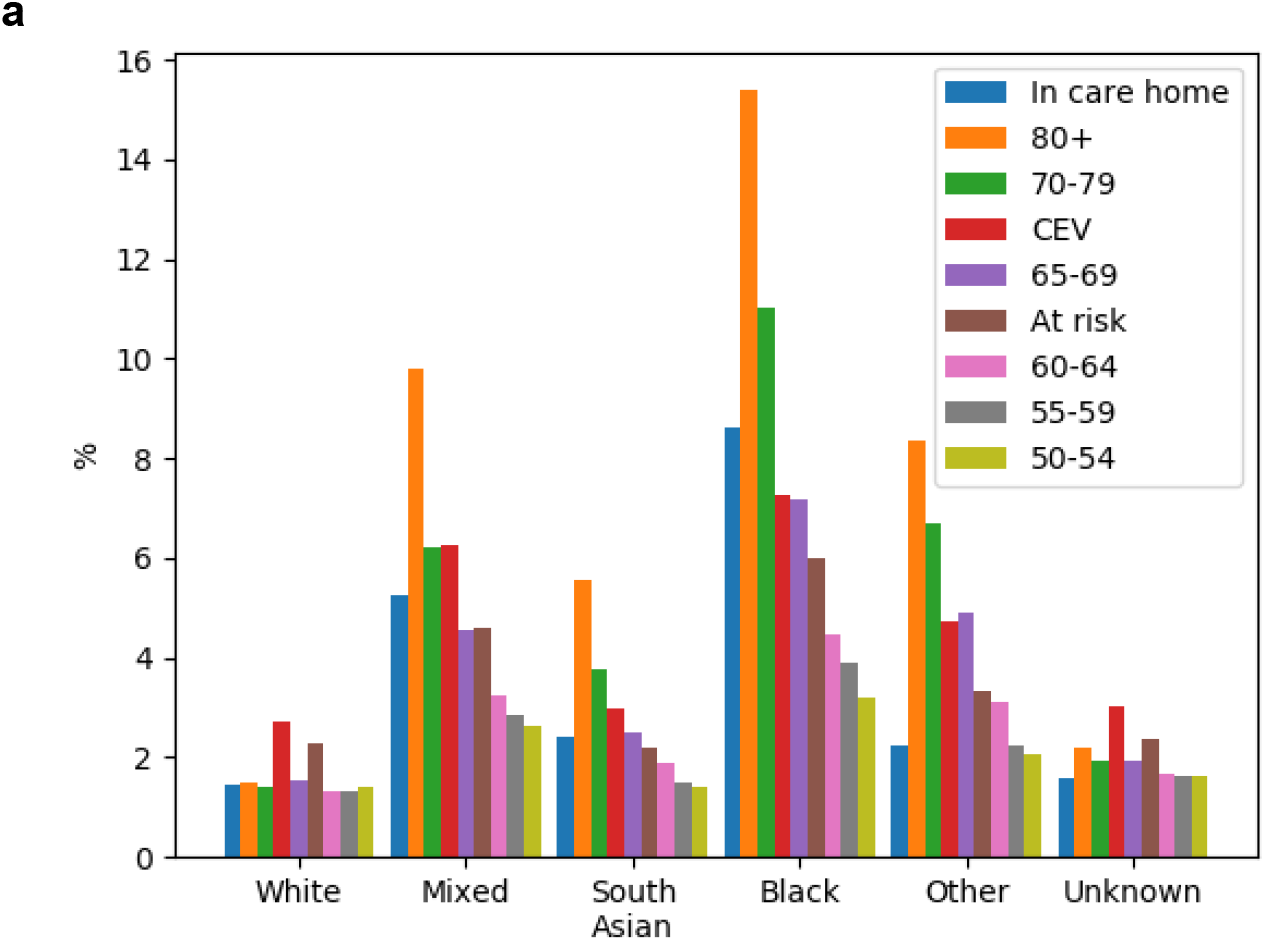
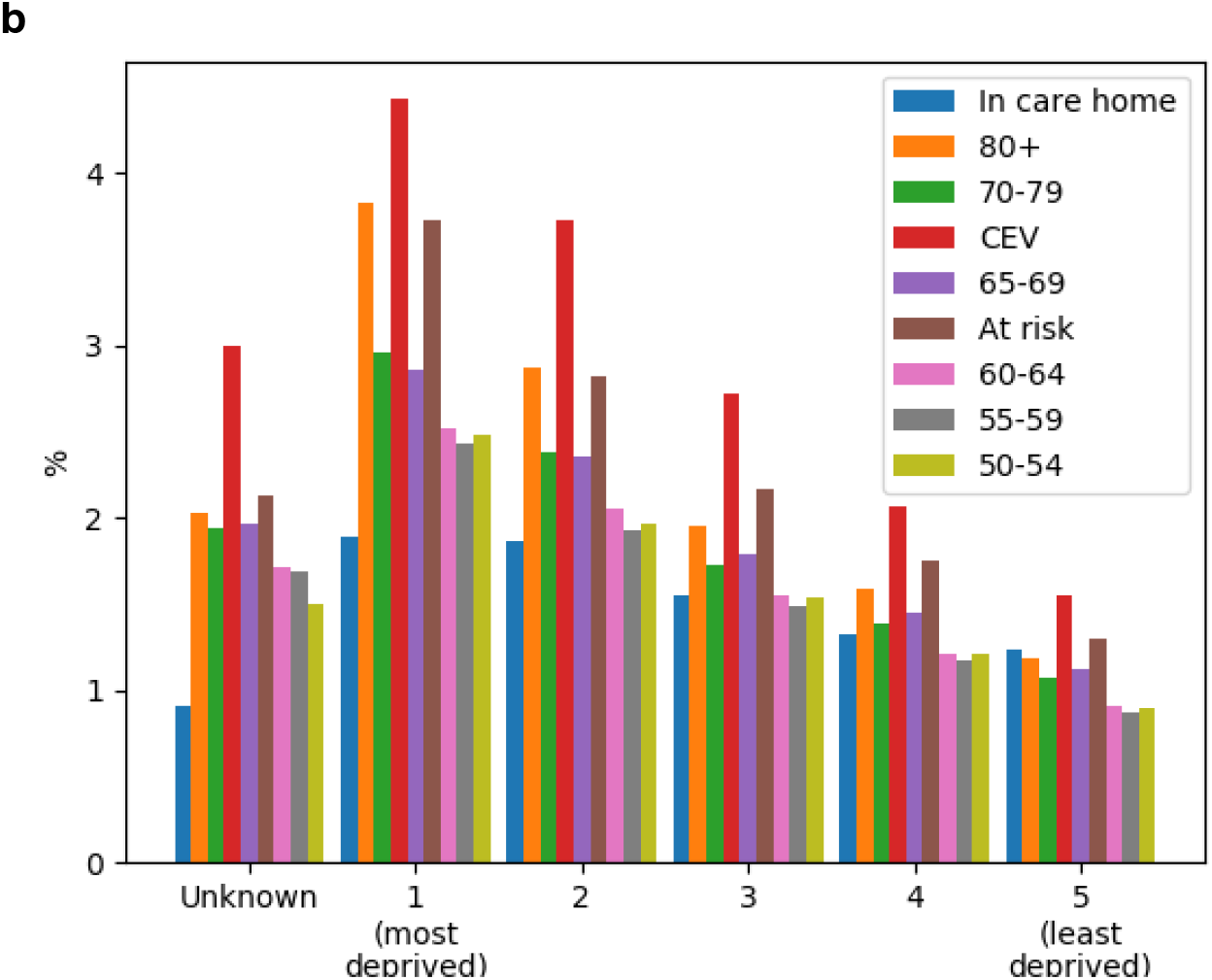
Percentage of those in each (a) ethnic group and (b) IMD band who had a decline recorded and are unvaccinated in OpenSAFELY as at 25th May 2021, split by by priority group.

**Figure 3.**
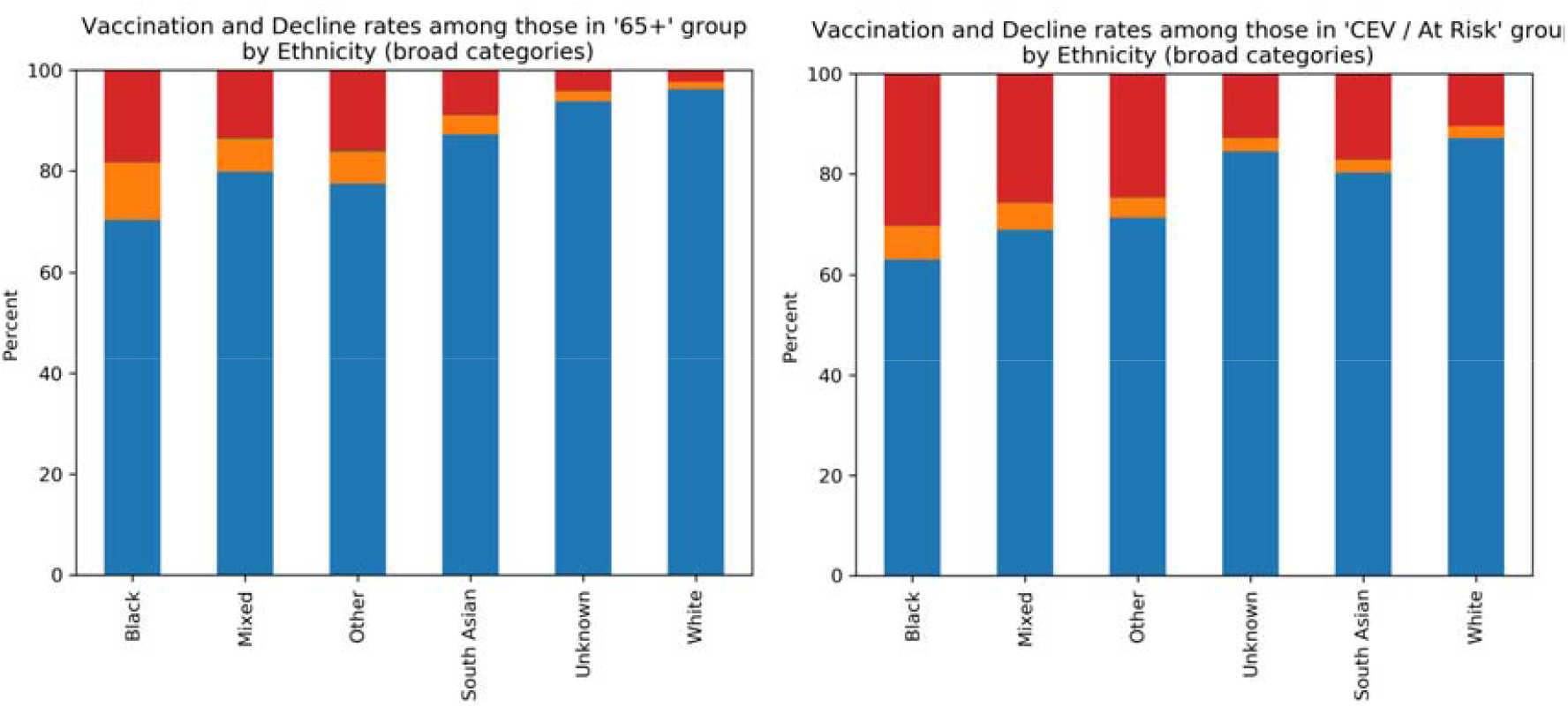
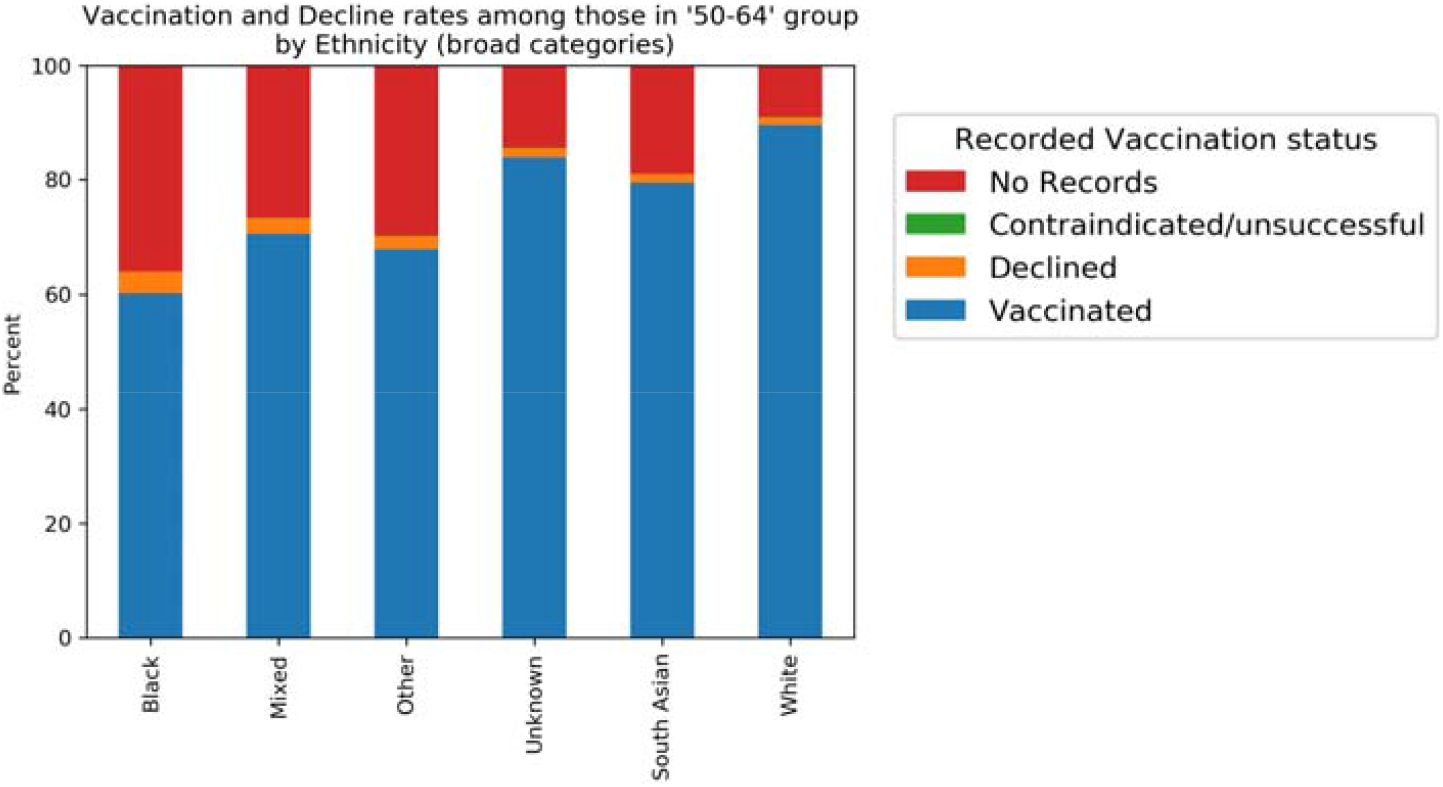
COVID-19 vaccination status recorded for patients in OpenSAFELY as at 25th May 2021 for combined priority groups 65+, CEV / At Risk, and 50-64, split by broad ethnic groups. “Declined” excludes patients with a recorded vaccination. “Vaccinated” includes those previously recorded as declining.

There was a clear trend towards increased recording of declines with increasing deprivation (least deprived quintile 0.9-1.5%, most deprived 2.4-4.4% excluding care homes; figure 2b). This was also consistently increasing over time (figure S1d-f). Presence of a Severe Mental Health condition was associated with lower vaccination rates and more declines being recorded, and a similar but less divergent pattern was seen in those with a Learning Disability (table S3a,b). Among all those with a recent pregnancy (only applicable in the CEV/At Risk group), vaccination rates were much lower compared with others of childbearing age (37.95% CEV/At Risk vs 67.06% and 72.56% for ages 16-29 and 30-39 respectively), more declines were recorded (5.94% vs 4.22% and 3.74%), and more had no vaccine-related records (55.65% vs 28.57% and 23.55% (table S3b).

### Patients who were recorded as declining and later had a vaccination

Of all those in priority groups who have had a decline recorded at any point, 18.9% later received a vaccination (Table 1). This conversion rate from “declined” to vaccinated ranged from 13.1% in the At Risk group to 30.7% in the 80+ group (figure 4a). This pattern was broadly similar in each ethnic group, but with the South Asian population generally higher, having the highest conversion rate in all but two priority groups (figure 4b). Among combined cohorts 65+, CEV/At risk, and 50-64, the conversion rates in the South Asian group were 32.3%, 25.2%, and 19.3% respectively, vs 22.8%,15.5%,and 16.8% for all other ethnicities combined. The time delay between the recorded decline and the first dose being received was primarily 0-2 weeks in the younger groups (aged 50-64), in contrast to the older groups which had a wider range of time delays.

**Figure 4.**
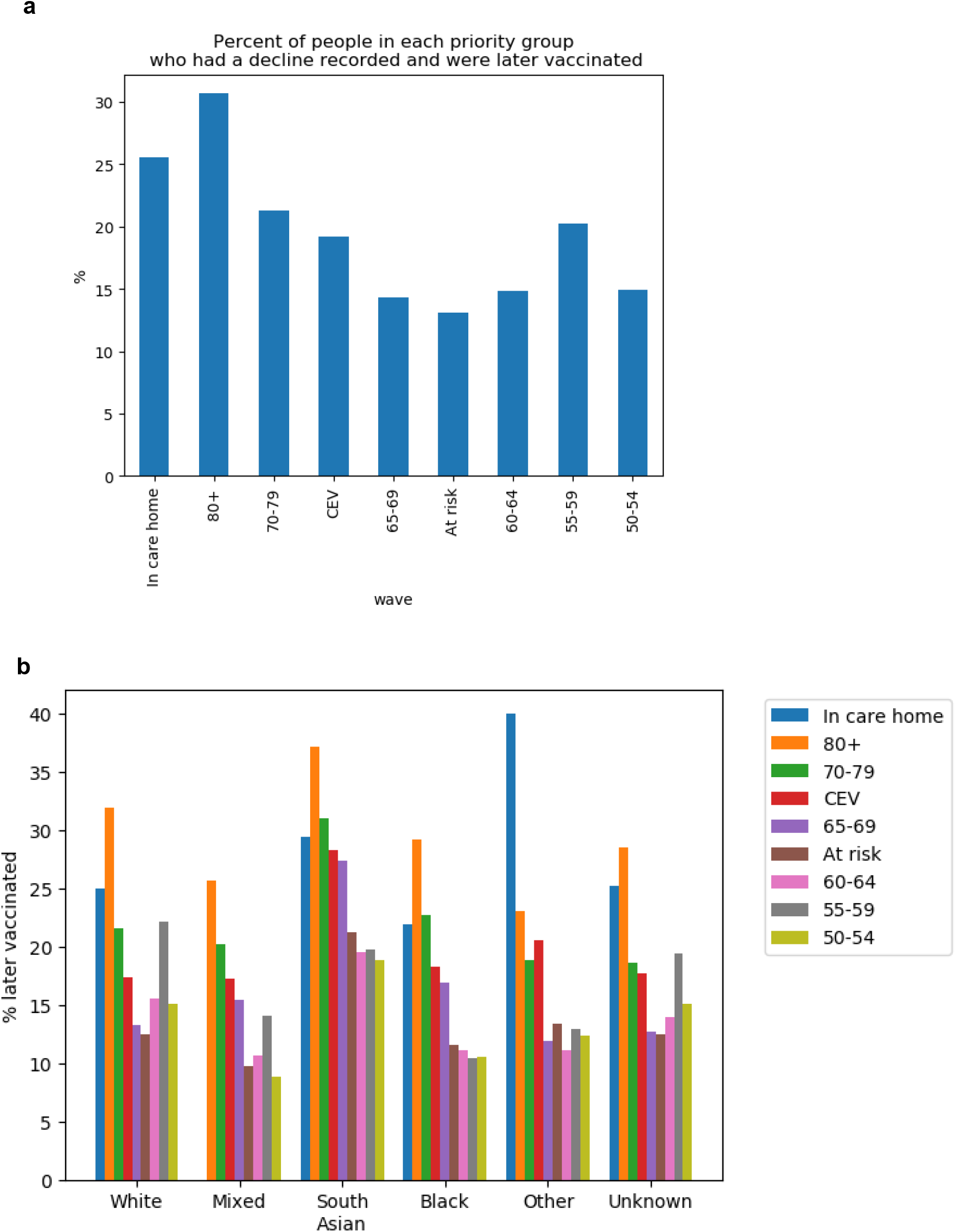
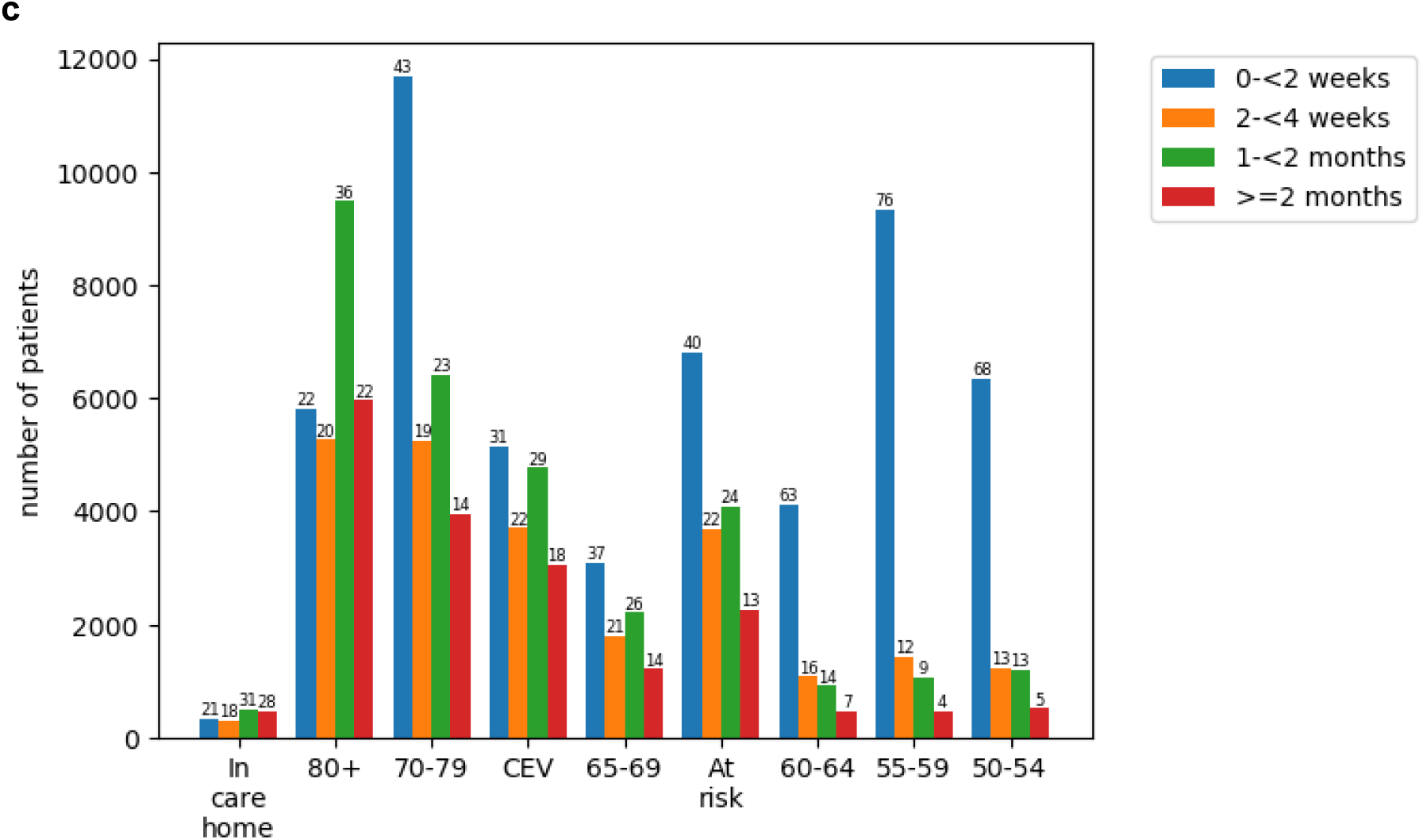
Patients in each priority group who were later vaccinated, after previously being recorded as declining a COVID-19 vaccine. (a) Percentage of those in each priority group who were later vaccinated; (b) Percentage of those in each priority group who were later vaccinated by ethnicity and priority group; (c) Number of patients in each priority group who were later vaccinated, by length of time between the recorded decline and the vaccination (labels show percentages). The youngest eligible group (50-54) was eligible for only one month at the time of analysis, though some healthcare workers counted in this group will have been eligible much longer.

### Variation by practice

Almost all practices (98.8%) had at least one patient recorded as declining the vaccine (6,290/6,364, limited to practices with >250 patients in priority groups and >10 vaccinated patients). There was a broad spread of rates per practice, with just over half of practices having <15 patients recorded as declining per 1000 registered patients, and most (90%) having 50 or fewer (Figure 5a). The majority of practices (90%) had 60 or fewer declines recorded per 1000 vaccinated patients (Figure 5b). However, there was a long tail with some practices having 300 or more recorded declines per 1000 vaccinated patients. Plotting against the number of priority group patients per practice indicates no strong correlation with practice size, though smaller practices were slightly more likely to have higher rates of declines recorded (Figure S2).

**Figure 5.**
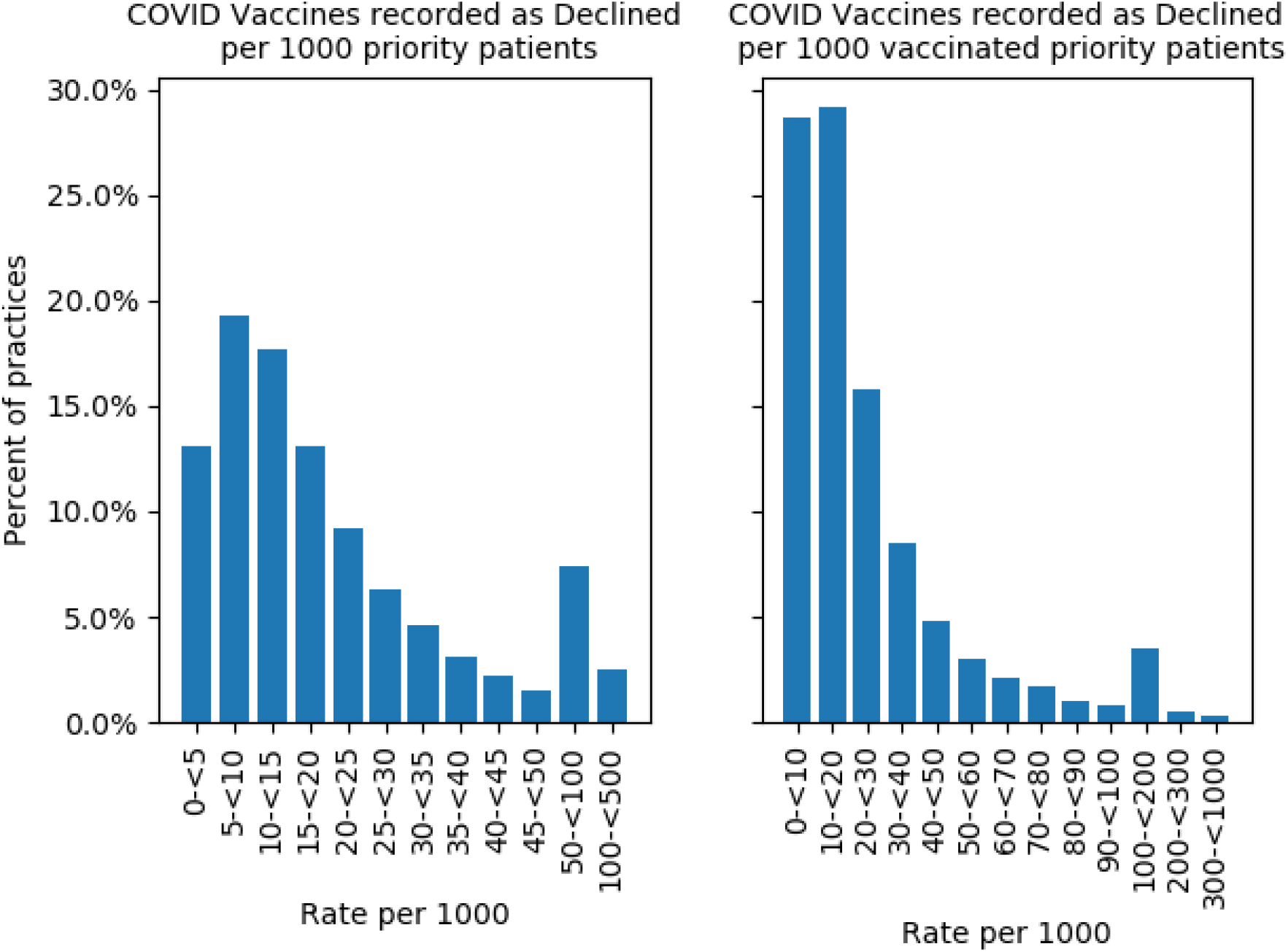
Histograms showing variation in the number of patients in priority groups per practice recorded as declining a COVID-19 vaccination (a) per 1000 patients in priority groups, and (b) per 1000 vaccinated patients in priority groups, as at 25th May 2021. Only includes unvaccinated patients. Practices with 250 or fewer registered patients in priority groups and those with 10 or fewer vaccinated patients were excluded.

## Discussion

### Summary

Overall, of currently registered people in priority groups almost half a million (2.0%) have been recorded as declining and remain unvaccinated, while 8.8% were unvaccinated without a recorded reason. Recorded declines were most common in the Clinically Extremely Vulnerable (CEV) group. Patients from ethnic minority groups and more deprived areas had higher rates of vaccine decline codes. Codes for declining COVID-19 vaccines were present in almost all practices, but there was substantial variation in rates. Of all those in priority groups who had a decline recorded at any point, 18.9% were later vaccinated.

### Strengths and weaknesses

The key strength of this study is its unprecedented scale: our source population includes 57.9 million people, over 95% of the population in England. This was achieved by developing and deploying data management and data analysis software inside the EHR vendors’ infrastructure, where the patient data already resides. Another key strength is that we identified patients in JCVI priority groups by directly implementing the full official SNOMED-CT codelists and logic for the national PRIMIS COVID-19 Vaccination Uptake Reporting Specification, thus ensuring that our cohorts are perfectly in line with national procedures and GP expectations.

We recognise some limitations with our analysis. Our population, though extremely large, may not be fully representative of the full eligible population: it does not include individuals not registered with a general practice, or the 4% of patients registered at practices not using TPP or EMIS. We include only currently registered and living patients, and exclude those who have moved away or died during the vaccination campaign. Primary care records, whilst detailed and longitudinal, cannot be used to determine vaccine eligibility through reasons of employment, and as such our priority groups which include working-age people will have a subset who were offered the vaccination earlier than others. As there is no national guidance on the use of decline codes it is likely that there is variation in how these codes are being used in practice. The rate of declines being recorded in each priority group is likely to be influenced by the length of time each has been eligible for the vaccine, and the number of times practices have attempted to contact them, highlighting the need for ongoing monitoring of this issue: for example the 80+ group were invited from the start of the campaign in December 2020, while the 50-54 group were invited from mid-March 2021 (NHS website 2021). It is not possible to determine whether those with no vaccine-related records are effectively declining by not responding to the invitations, are intending to be vaccinated later, or have failed to be reached (e.g. outdated/incorrect contact details or have left the country). It is possible that some have not been approached at all, although all patients in priority groups had reportedly been offered the vaccine just over one month prior to our latest data (NHS England 2021), therefore this should be largely restricted to those without valid contact details. Data flow from mass vaccination centres is thought to be largely complete, but some vaccination records may have been missed, for example vaccines delivered in inpatient settings; this may disproportionately affect the CEV and At Risk groups, for example those with kidney disease (Kidney Care UK 2021). Finally, some patients with another code, e.g. “did not attend” (DNA), may also be declining but not been recorded as such. DNAs are likely to be vastly underrepresented in our data as data flowing from mass vaccination centres only includes successful vaccinations, therefore DNAs will only be recorded for appointments booked in surgeries.

### Findings in Context

Among priority groups, the proportion of people recorded as declining and being unvaccinated was highest in the CEV and At Risk groups, even when comparing individual 5-year age bands. In a 2020 survey, CEV (shielding) status was associated with lower self-reported COVID-19 vaccine hesitancy (Batty, Deary, and Altschul 2021). However, we also found the percentage who were vaccinated was higher in the CEV/At Risk groups compared to others of the same age. Therefore, a likely explanation for our finding is that those at increased risk due to their clinical conditions were sent more follow-up invitations, giving a greater opportunity for a vaccine to be administered or a decline to be recorded.

Our finding of higher rates of declines being recorded in Black and South Asian groups is generally consistent with survey data on intention to accept the COVID-19 vaccine (Parveen, Mohdin, and McIntyre 2021; Robinson, Jones, and Daly 2020; Allington et al. 2021; Royal Society of Public Health 2020; Robertson et al. 2021), and with previous research on variation in vaccine coverage in other vaccination campaigns historically (Ward et al. 2017; Loiacono et al. 2020; Jain et al. 2018).

We found that 18.9% of those initially recorded as declining were later vaccinated. As well as those who genuinely changed their mind, this will include some who always intended to be vaccinated, such as those who temporarily declined (eg. due to illness), rejected a repeated invitation after already booking a vaccine, or had a decline recorded in error or for administrative reasons. It may also reflect changing preferences over time: a recent survey indicated that 52% of those reporting they would definitely not have the vaccine in Nov/Dec 2020 have since accepted it when offered (and 15% of those not yet offered were likely to accept) (Allington et al. 2021), while another survey noted a reduction in hesitancy from 26·9% in October 2020 to 16·9% in Jan/Feb 2021 (Freeman et al. 2021), indicating substantial shifts in people’s preferences as the campaign has progressed. We found the conversion rate was broadly similar across ethnic groups (but slightly higher in the South Asian population).

### Policy Implications and further research

Almost all practices had at least one patient with a declined code, indicating that these codes are widely used. However, there was substantial variation between practices, which could reflect some differences in administrative processes around the application of these codes as well as variation in patient preference in different localities. Due to the speed and scale of the vaccine roll-out, development of SNOMED-CT codes related to COVID-19 vaccines was, understandably, restricted to cover core clinical information about an individual’s immunisation that could be shared to inform any subsequent clinical decisions. Unlike other mass vaccination campaigns in England, many of the codes related to a patient’s administrative journey, the “call and recall” system, were not developed (NHS Digital 2021). Development of further administrative codes specific to the COVID-19 vaccination should be considered to facilitate future “booster” campaigns. Until then, the limited number of available decline codes will inevitably be used in a broad range of individual patient situations, as such we recommend detailed survey and/or qualitative research with patients and NHS staff should be conducted to provide more descriptive information on how these codes are being used and shed light on the differences between groups. The South Asian group may be a priority group for such research in order to identify the reasons for the high prevalence of a decline code followed by subsequent vaccination.

### Summary

Clinical codes indicative of COVID-19 vaccinations being declined are widely recorded in English general practice, and more common in patients from deprived areas and Black and South Asian groups. The reasons for this require further exploration, we suggest questionnaire and qualitative work, including among those who go on to subsequently receive a vaccine.

## Supporting information

STROBE checklist

## Data Availability

Data was accessed through the OpenSAFELY platform, as per Supplementary Materials. Data management and analysis was performed using the OpenSAFELY software libraries and Python, both implemented using Python3. All code for data management and analysis for this paper is shared for scientific review and re-use under open licenses on GitHub (https://github.com/opensafely/covid-vaccination-not-received). All codelists are available for inspection and re-use from https://codelists.opensafely.org. All code for the OpenSAFELY platform for data management, analysis and secure code execution is shared for review and re-use under open licenses at GitHub.com/OpenSAFELY.

https://github.com/opensafely/covid-vaccine-not-received

## Administrative

## Acknowledgements

We are very grateful for all the support received from the EMIS and TPP Technical Operations team throughout this work, and for generous assistance from the information governance and database teams at NHS England / NHSX.

## Information governance and ethical approval

NHS England is the data controller; EMIS and TPP are the data processors; and the key researchers on OpenSAFELY are acting on behalf of NHS England. This implementation of OpenSAFELY is hosted within the EMIS and TPP environments which are accredited to the ISO 27001 information security standard and are NHS IG Toolkit compliant;(“BETA – Data Security Standards - NHS Digital” n.d., “Data Security and Protection Toolkit - NHS Digital” n.d.) patient data has been pseudonymised for analysis and linkage using industry standard cryptographic hashing techniques; all pseudonymised datasets transmitted for linkage onto OpenSAFELY are encrypted; access to the platform is via a virtual private network (VPN) connection, restricted to a small group of researchers; the researchers hold contracts with NHS England and only access the platform to initiate database queries and statistical models; all database activity is logged; only aggregate statistical outputs leave the platform environment following best practice for anonymisation of results such as statistical disclosure control for low cell counts.(“ISB1523: Anonymisation Standard for Publishing Health and Social Care Data - NHS Digital” n.d.) The OpenSAFELY research platform adheres to the obligations of the UK General Data Protection Regulation (GDPR) and the Data Protection Act 2018. In March 2020, the Secretary of State for Health and Social Care used powers under the UK Health Service (Control of Patient Information) Regulations 2002 (COPI) to require organisations to process confidential patient information for the purposes of protecting public health, providing healthcare services to the public and monitoring and managing the COVID-19 outbreak and incidents of exposure; this sets aside the requirement for patient consent.(Secretary of State for Health and Social Care - UK Government 2020) Taken together, these provide the legal bases to link patient datasets on the OpenSAFELY platform. GP practices, from which the primary care data are obtained, are required to share relevant health information to support the public health response to the pandemic, and have been informed of the OpenSAFELY analytics platform.

This study was approved by the Health Research Authority (REC reference 20/LO/0651) and by the LSHTM Ethics Board (reference 21863).

### Guarantor

BG/LS are guarantors of the OpenSAFELY project.

## References

Allington, Daniel, Bobby Duffy, Vivienne Moxham-Hall, Siobhan McAndrew, and George Murkin. 2021. “Covid-19: Vaccine Take-up and Trust.” June 12, 2021. https://www.kcl.ac.uk/policy-institute/assets/covid-19-vaccine-take-up-and-trust.pdf.

Batty, G. David, Ian J. Deary, and Drew Altschul. 2021. “Pre-Pandemic Mental and Physical Health as Predictors of COVID-19 Vaccine Hesitancy: Evidence from a UK-Wide Cohort Study.” medRxiv : The Preprint Server for Health Sciences, April. https://doi.org/10.1101/2021.04.27.21256185.

“BETA – Data Security Standards - NHS Digital.” n.d. NHS Digital. Accessed April 30, 2020. https://digital.nhs.uk/about-nhs-digital/our-work/nhs-digital-data-and-technology-standards/framework/beta---data-security-standards.

“COVID-19 Vaccination Uptake Reporting Specification.” n.d. PRMIS. Accessed March 18, 2021. https://www.nottingham.ac.uk/primis/covid-19/covid-19.aspx.

“Data Security and Protection Toolkit - NHS Digital.” n.d. NHS Digital. Accessed April 30, 2020. https://digital.nhs.uk/data-and-information/looking-after-information/data-security-and-information-governance/data-security-and-protection-toolkit.

Freeman, Daniel, Bao Sheng Loe, Ly-Mee Yu, Jason Freeman, Andrew Chadwick, Cristian Vaccari, Milensu Shanyinde, et al. 2021. “Effects of Different Types of Written Vaccination Information on COVID-19 Vaccine Hesitancy in the UK (OCEANS-III): A Single-Blind, Parallel-Group, Randomised Controlled Trial.” The Lancet. Public Health 6 (6): e416–27.

“ISB1523: Anonymisation Standard for Publishing Health and Social Care Data - NHS Digital.” n.d. NHS Digital. Accessed April 30, 2020. https://digital.nhs.uk/data-and-information/information-standards/information-standards-and-data-collections-including-extractions/publications-and-notifications/standards-and-collections/isb1523-anonymisation-standard-for-publishing-health-and-social-care-data.

Jain, Anu, Jemma L. Walker, Rohini Mathur, Harriet J. Forbes, Sinéad M. Langan, Liam Smeeth, Albert J. van Hoek, and Sara L. Thomas. 2018. “Zoster Vaccination Inequalities: A Population Based Cohort Study Using Linked Data from the UK Clinical Practice Research Datalink.” PloS One 13 (11): e0207183.

Loiacono, Matthew M., Salaheddin M. Mahmud, Ayman Chit, Robertus van Aalst, Jeffrey C. Kwong, Nicholas Mitsakakis, Luke Skinner, Edward Thommes, Hélène Bricout, and Paul Grootendorst. 2020. “Patient and Practice Level Factors Associated with Seasonal Influenza Vaccine Uptake among at-Risk Adults in England, 2011 to 2016: An Age-Stratified Retrospective Cohort Study.” Vaccine: X 4 (April): 100054.

MacKenna, Brian. 2019. “What Is the Dm+d? The NHS Dictionary of Medicines and Devices.” EBM DataLab. August 27, 2019. https://ebmdatalab.net/what-is-the-dmd-the-nhs-dictionary-of-medicines-and-devices/.

NHS Digital. 2021. “Delen: Home : Terminology & Classifications News : COVID-19 Vaccination Codes.” NHS Digital. March 22, 2021. https://hscic.kahootz.com/t_c_home/viewBlogArticle?articleID=655513.

NHS-England. 2021. “COVID-19 Vaccine Programme: Maximising Vaccine Uptake in Underserved Communities.” March 2021. https://www.england.nhs.uk/coronavirus/wp-content/uploads/sites/52/2021/03/C1226-maximising-vaccine-uptake-in-underserved-communities-a-framework-.pdf.

NHS England. 2021. “Next Phase of NHS COVID-19 Vaccination Campaign.” April 13, 2021. https://www.england.nhs.uk/coronavirus/wp-content/uploads/sites/52/2021/04/C1252-next-phase-of-nhs-covid-19-vaccination-campaign.pdf.

Parveen, Nazia, Aamna Mohdin, and Niamh McIntyre. 2021. “Call to Prioritise Minority Ethnic Groups for Covid Vaccines.” The Guardian, January 18, 2021. http://www.theguardian.com/world/2021/jan/18/call-to-prioritise-minority-ethnic-groups-for-covid-vaccines.

Public Health England. 2020. “The Green Book, Chapter 14a - COVID-19.” GOV.UK. November 27, 2020. https://www.gov.uk/government/publications/covid-19-the-green-book-chapter-14a.

Robertson, Elaine, Kelly S. Reeve, Claire L. Niedzwiedz, Jamie Moore, Margaret Blake, Michael Green, Srinivasa Vittal Katikireddi, and Michaela J. Benzeval. 2021. “Predictors of COVID-19 Vaccine Hesitancy in the UK Household Longitudinal Study.” Brain, Behavior, and Immunity 94 (May): 41–50.

Robinson, Eric, Andrew Jones, and Michael Daly. 2020. “International Estimates of Intended Uptake and Refusal of COVID-19 Vaccines: A Rapid Systematic Review and Meta-Analysis of Large Nationally Representative Samples.” medRxiv. https://www.medrxiv.org/content/10.1101/2020.12.01.20241729v1.abstract.

Royal Society of Public Health. 2020. “New Poll Finds BAME Groups Less Likely to Want COVID Vaccine.” December 16, 2020. https://www.rsph.org.uk/about-us/news/new-poll-finds-bame-groups-less-likely-to-want-covid-vaccine.html.

Secretary of State for Health and Social Care - UK Government. 2020. “Coronavirus (COVID-19): Notification to Organisations to Share Information.” April 1, 2020. https://web.archive.org/web/20200421171727/ https://www.gov.uk/government/publications/coronavirus-covid-19-notification-of-data-controllers-to-share-information.

The OpenSAFELY Collaborative, Helen J. Curtis, Peter Inglesby, Caroline E. Morton, Brian MacKenna, Alex J. Walker, Jessica Morley, et al. 2021. “Trends and Clinical Characteristics of COVID-19 Vaccine Recipients: A Federated Analysis of 57.9 Million Patients’ Primary Care Records in Situ Using OpenSAFELY.” bioRxiv. medRxiv. https://doi.org/10.1101/2021.01.25.21250356.

Ward, Charlotte, Lisa Byrne, Joanne M. White, Gayatri Amirthalingam, Karen Tiley, and Michael Edelstein. 2017. “Sociodemographic Predictors of Variation in Coverage of the National Shingles Vaccination Programme in England, 2014/15.” Vaccine 35 (18): 2372–78.

