## Supplementary Information for "Recording of “COVID-19 vaccine declined” among vaccination priority groups: a cohort study on 57.9 million NHS patients’ primary care records in situ using OpenSAFELY"

### Appendix

***Table S1. Classification of SNOMED codes related to COVID-19 vaccines being given.*** *These clinical codes will reflect only a small minority of cases, as vaccines are almost always recorded according to the product given.*

| SNOMED Code | Description | Codelist |
|---|---|---|
| 90640007 | Coronavirus vaccination | COV-19 vacc given |
| 840534001 | Severe acute respiratory syndrome coronavirus 2 vaccination | covadm1 |
| 1324671000000103 | Immunisation course to achieve immunity against SARS-CoV-2 (severe acute respiratory syndrome coronavirus 2) | COV-19 Vacc given |
| 1324681000000101 | Administration of first dose of SARS-CoV-2 (severe acute respiratory syndrome coronavirus 2) vaccine | Covadm1<br>COV-19 vacc given |
| 1324691000000104 | Administration of second dose of SARS-CoV-2 (severe acute respiratory syndrome coronavirus 2) vaccine | Covadm2<br>COV-19 vacc given |
| 1324851000000106 | SARS-CoV-2 (severe acute respiratory syndrome coronavirus 2) immunisation course started | COV-19 vacc given |

***Sources: COVID-19 Vaccination Codes*** *(NHS Digital 2021); National COVID-19 Vaccination Uptake Reporting Specification ("COVID-19 Vaccination Uptake Reporting Specification" n.d.)*

***Table S2. Classification of SNOMED codes related to COVID-19 vaccines being (a) declined or (b) otherwise not done.*** *"Not done" codes are only counted for patients with no record of any dose being given.*

| Classification | SNOMED Code | Description | Codelist(s) |
|---|---|---|---|
| **(a) Declined** | 1324741000000101 | SARS-CoV-2 (severe acute respiratory syndrome coronavirus 2) vaccination first dose declined | 1st dose declined |
| | 1324811000000107 | SARS-CoV-2 (severe acute respiratory syndrome coronavirus 2) immunisation course declined | 1st dose declined<br>2nd dose declined |
| | 1324721000000108 | SARS-CoV-2 (severe acute respiratory syndrome coronavirus 2) vaccination dose declined | 1st dose declined<br>2nd dose declined |
| | 1240651000000109 | *[inactive]* SARS-CoV-2 (severe acute respiratory syndrome coronavirus 2) vaccination declined | 1st dose declined<br>2nd dose declined |
| | 1324751000000103 | SARS-CoV-2 (severe acute respiratory syndrome coronavirus 2) vaccination second dose declined | 2nd dose declined |
| **(b) Not done** | 1240631000000102 | Did not attend SARS-CoV-2 vaccination | 1st appt DNA<br>2nd appt DNA |
| | 1324831000000104 | Did not attend for first dose of SARS-CoV-2 (severe acute respiratory syndrome coronavirus 2) vaccine | 1st appt DNA |
| | 1324841000000108 | Did not attend for second dose of SARS-CoV-2 (severe acute respiratory syndrome coronavirus 2) vaccine | 2nd appt DNA |

| | | | |
|---|---|---|---|
| | 1324661000000105 | Adverse reaction to SARS-CoV-2 (severe acute respiratory syndrome coronavirus 2) vaccine | allergy/contra |
| | 1324711000000102 | Allergy to SARS-CoV-2 (severe acute respiratory syndrome coronavirus 2) vaccine | allergy/contra |
| | 1324731000000105 | SARS-CoV-2 (severe acute respiratory syndrome coronavirus 2) immunisation course not indicated | allergy/contra |
| | 1324761000000100 | SARS-CoV-2 (severe acute respiratory syndrome coronavirus 2) immunisation course contraindicated | allergy/contra |
| | 1240661000000107 | *[inactive]* SARS-CoV-2 (severe acute respiratory syndrome coronavirus 2) vaccination contraindicated | allergy/contra |
| | 1240671000000100 | *[inactive]* SARS-CoV-2 (severe acute respiratory syndrome coronavirus 2) vaccination not indicated | allergy/contra |
| | 1324821000000101 | SARS-CoV-2 (severe acute respiratory syndrome coronavirus 2) immunisation course not done | 1st dose not given |
| | 1240681000000103 | *[inactive]* Severe acute respiratory syndrome coronavirus 2 vaccination not done | 1st dose not given |
| | 1324771000000107 | SARS-CoV-2 (severe acute respiratory syndrome coronavirus 2) vaccination dose not given | 1st dose not given |
| | 1324781000000109 | SARS-CoV-2 (severe acute respiratory syndrome coronavirus 2) vaccination first dose not given | 1st dose not given |
| | 1324791000000106 | SARS-CoV-2 (severe acute respiratory syndrome coronavirus 2) vaccination second dose not given | 2nd dose not given |
| | 1324861000000109 | SARS-CoV-2 (severe acute respiratory syndrome coronavirus 2) immunisation course abandoned | 2nd dose not given |
| | 1240701000000101 | Severe acute respiratory syndrome coronavirus 2 vaccine not available | COV19 vacc unavailable |

***Source:* *COVID-19 Vaccination Codes*** *(NHS Digital 2021); plus three additional codes were identified in the national COVID-19 Vaccination Uptake Reporting Specification ("COVID-19 Vaccination Uptake Reporting Specification" n.d.), or in a SNOMED browser. Inactive codes indicate those which are no longer available for use but may have been entered in patient records previously.*

***Table S3***

***Percentage of population in each combined priority group [(a) 65+ (b) CEV/At Risk, (c) 50-64] who are recorded as vaccinated, declined and unvaccinated, contraindicated/unsuccessful, or with no vaccine records, according to demographic features, as at 25 May 2021.*** *Percentages may not sum to 100 due to rounding. Patient counts rounded to the nearest 7.*

***Table 3a. Ages 65+ (including care home residents)***

| **Category** | **Group** | **Total in 65+ cohort** | **Vaccinated (% of total)** | **Declined-Unvaccinated (% of total)** | **Contraindicated/ unsuccessful (% of total)** | **No Records (% of total)** |
|---|---|---|---|---|---|---|
| Age Band | 65-<70 | 2,516,955 | 2,324,469 (92.35%) | 46,018 (1.83%) | 1,463 (0.06%) | 145,005 (5.76%) |
| | 70-<75 | 2,783,242 | 2,640,092 (94.86%) | 51,107 (1.84%) | 1,610 (0.06%) | 90,433 (3.25%) |
| | 75-<80 | 2,023,665 | 1,939,441 (95.84%) | 34,244 (1.69%) | 882 (0.04%) | 49,098 (2.43%) |
| | 80-<85 | 1,408,400 | 1,352,071 (96.00%) | 27,048 (1.92%) | 532 (0.04%) | 28,749 (2.04%) |
| | 85-<120 | 1,357,335 | 1,295,980 (95.48%) | 29,176 (2.15%) | 504 (0.04%) | 31,675 (2.33%) |
| Sex | Female | 5,445,440 | 5,167,911 (94.90%) | 107,359 (1.97%) | 2,401 (0.04%) | 167,769 (3.08%) |
| | Male | 4,644,171 | 4,384,163 (94.40%) | 80,262 (1.73%) | 2,562 (0.06%) | 177,184 (3.82%) |
| High Level Ethnicity | White | 6,384,245 | 6,140,638 (96.18%) | 92,659 (1.45%) | 2,219 (0.03%) | 148,729 (2.33%) |
| | Mixed | 40,173 | 32,081 (79.86%) | 2,597 (6.46%) | 70 (0.17%) | 5,425 (13.50%) |
| | South Asian | 310,177 | 270,592 (87.24%) | 11,564 (3.73%) | 273 (0.09%) | 27,748 (8.95%) |
| | Black | 119,616 | 84,245 (70.43%) | 13,349 (11.16%) | 175 (0.15%) | 21,847 (18.26%) |
| | Other | 62,559 | 48,440 (77.43%) | 3,934 (6.29%) | 126 (0.20%) | 10,059 (16.08%) |
| | Unknown | 3,172,813 | 2,976,050 (93.80%) | 63,490 (2.00%) | 2,135 (0.07%) | 131,138 (4.13%) |
| IMD Band | Unknown | 82,649 | 78,309 (94.75%) | 1,617 (1.96%) | 49 (0.06%) | 2,674 (3.24%) |
| | 1 (most deprived) | 1,418,151 | 1,292,823 (91.16%) | 44,093 (3.11%) | 1,029 (0.07%) | 80,206 (5.66%) |
| | 2 | 1,750,252 | 1,627,276 (92.97%) | 43,393 (2.48%) | 840 (0.05%) | 78,743 (4.50%) |
| | 3 | 2,142,014 | 2,033,731 (94.94%) | 38,437 (1.79%) | 952 (0.04%) | 68,894 (3.22%) |
| | 4 | 2,298,051 | 2,203,257 (95.88%) | 33,313 (1.45%) | 1,029 (0.04%) | 60,452 (2.63%) |
| | 5 (least deprived) | 2,398,466 | 2,316,650 (96.59%) | 26,740 (1.11%) | 1,106 (0.05%) | 53,970 (2.25%) |
| Severe Mental Health | no | 9,989,966 | 9,462,292 (94.72%) | 183,589 (1.84%) | 4,907 (0.05%) | 339,178 (3.40%) |
| | yes | 99,638 | 89,782 (90.11%) | 4,025 (4.04%) | 63 (0.06%) | 5,768 (5.79%) |
| Learning Disability | no | 10,065,412 | 9,529,114 (94.67%) | 187,089 (1.86%) | 4,956 (0.05%) | 344,253 (3.42%) |
| | yes | 24,192 | 22,953 (94.88%) | 525 (2.17%) | 21 (0.09%) | 693 (2.86%) |

***Table S3b. CEV/At Risk.*** *CEV includes ages 16-69, At Risk includes ages 16-64. Pregnant/recent pregnancy includes those with a pregnancy code in the 8 months prior to 25 May.*

| Category | Group | Total in CEV/At Risk cohort | Vaccinated (% of total) | Declined-Unvaccinated (% of total) | Contraindicated / unsuccessful (% of total) | No Records (% of total) |
|---|---|---|---|---|---|---|
| Age Band | 16-<30 | 664,272 | 445,431 (67.06%) | 28,014 (4.22%) | 1,071 (0.16%) | 189,756 (28.57%) |
| | 30-<40 | 921,655 | 668,780 (72.56%) | 34,496 (3.74%) | 1,365 (0.15%) | 217,014 (23.55%) |
| | 40-<50 | 1,265,278 | 1,053,220 (83.24%) | 35,203 (2.78%) | 1,106 (0.09%) | 175,749 (13.89%) |
| | 50-<55 | 919,177 | 814,443 (88.61%) | 22,043 (2.40%) | 602 (0.07%) | 82,089 (8.93%) |
| | 55-<60 | 1,106,588 | 1,003,954 (90.73%) | 23,471 (2.12%) | 630 (0.06%) | 78,533 (7.10%) |
| | 60-<65 | 1,167,922 | 1,082,123 (92.65%) | 22,015 (1.88%) | 553 (0.05%) | 63,231 (5.41%) |
| | 65-<70 | 271,299 | 256,375 (94.50%) | 5,411 (1.99%) | 126 (0.05%) | 9,387 (3.46%) |
| Sex | Female | 3,139,913 | 2,656,472 (84.60%) | 86,289 (2.75%) | 2,695 (0.09%) | 394,457 (12.56%) |
| | Male | 3,176,292 | 2,667,875 (83.99%) | 84,392 (2.66%) | 2,723 (0.09%) | 421,302 (13.26%) |
| High Level Ethnicity | White | 3,553,998 | 3,095,582 (87.10%) | 85,603 (2.41%) | 2,947 (0.08%) | 369,866 (10.41%) |
| | Mixed | 89,950 | 62,062 (69.00%) | 4,760 (5.29%) | 140 (0.16%) | 22,988 (25.56%) |
| | South Asian | 572,208 | 459,228 (80.26%) | 14,609 (2.55%) | 483 (0.08%) | 97,888 (17.11%) |
| | Black | 260,589 | 164,234 (63.02%) | 17,402 (6.68%) | 308 (0.12%) | 78,645 (30.18%) |
| | Other | 90,370 | 64,575 (71.46%) | 3,521 (3.90%) | 133 (0.15%) | 22,141 (24.50%) |
| | Unknown | 1,749,083 | 1,478,645 (84.54%) | 44,765 (2.56%) | 1,428 (0.08%) | 224,245 (12.82%) |
| IMD Band | Unknown | 62,055 | 52,990 (85.39%) | 1,498 (2.41%) | 84 (0.14%) | 7,483 (12.06%) |
| | 1 (most deprived) | 1,614,564 | 1,256,983 (77.85%) | 64,393 (3.99%) | 1,715 (0.11%) | 291,473 (18.05%) |
| | 2 | 1,396,969 | 1,142,288 (81.77%) | 43,771 (3.13%) | 1,078 (0.08%) | 209,832 (15.02%) |
| | 3 | 1,214,472 | 1,047,578 (86.26%) | 28,357 (2.33%) | 1,015 (0.08%) | 137,522 (11.32%) |
| | 4 | 1,066,555 | 949,151 (88.99%) | 19,516 (1.83%) | 903 (0.08%) | 96,985 (9.09%) |
| | 5 (least deprived) | 961,576 | 875,350 (91.03%) | 13,111 (1.36%) | 665 (0.07%) | 72,450 (7.53%) |
| Pregnant / Recent pregnancy | no | 6,255,893 | 5,301,464 (84.74%) | 167,097 (2.67%) | 5,138 (0.08%) | 782,194 (12.50%) |
| | yes | 60,319 | 22,890 (37.95%) | 3,584 (5.94%) | 280 (0.46%) | 33,565 (55.65%) |
| Severe Mental Health | no | 5,912,137 | 5,038,740 (85.23%) | 150,031 (2.54%) | 4,858 (0.08%) | 718,508 (12.15%) |
| | yes | 404,068 | 285,614 (70.68%) | 20,650 (5.11%) | 553 (0.14%) | 97,251 (24.07%) |
| Learning Disability | no | 6,064,674 | 5,113,087 (84.31%) | 160,937 (2.65%) | 5,166 (0.09%) | 785,484 (12.95%) |
| | yes | 251,538 | 211,267 (83.99%) | 9,744 (3.87%) | 252 (0.10%) | 30,275 (12.04%) |

***Table S3c. Ages 50-64.*** *Those with severe mental health conditions or learning disability within these age bands are, by definition, included within the CEV/At Risk group (Table S3b above).*

| Category | Group | Total in 50-64 cohort | Vaccinated (% of total) | Declined-Unvaccinated (% of total) | Contraindicated / unsuccessful (% of total) | No Records (% of total) |
|---|---|---|---|---|---|---|
| Age Band | 50-<55 | 3,114,629 | 2,605,316 (83.65%) | 48,972 (1.57%) | 1,855 (0.06%) | 458,486 (14.72%) |
| | 55-<60 | 2,804,333 | 2,425,836 (86.50%) | 42,392 (1.51%) | 1,610 (0.06%) | 334,495 (11.93%) |
| | 60-<65 | 2,152,038 | 1,911,868 (88.84%) | 33,558 (1.56%) | 1,267 (0.06%) | 205,345 (9.54%) |
| Sex | Female | 4,058,264 | 3,604,398 (88.82%) | 58,457 (1.44%) | 1,645 (0.04%) | 393,764 (9.70%) |
| | Male | 4,012,750 | 3,338,629 (83.20%) | 66,465 (1.66%) | 3,094 (0.08%) | 604,562 (15.07%) |
| High Level Ethnicity | White | 4,687,473 | 4,198,481 (89.57%) | 63,665 (1.36%) | 1,876 (0.04%) | 423,451 (9.03%) |
| | Mixed | 75,481 | 53,305 (70.62%) | 2,135 (2.83%) | 63 (0.08%) | 19,978 (26.47%) |
| | South Asian | 327,831 | 260,386 (79.43%) | 5,089 (1.55%) | 140 (0.04%) | 62,216 (18.98%) |
| | Black | 216,104 | 130,256 (60.27%) | 8,043 (3.72%) | 154 (0.07%) | 77,651 (35.93%) |
| | Other | 109,725 | 74,515 (67.91%) | 2,583 (2.35%) | 105 (0.10%) | 32,522 (29.64%) |
| | Unknown | 2,654,365 | 2,226,063 (83.86%) | 43,393 (1.63%) | 2,408 (0.09%) | 382,501 (14.41%) |
| IMD Band | Unknown | 69,797 | 61,299 (87.82%) | 1,141 (1.63%) | 35 (0.05%) | 7,322 (10.49%) |
| | 1 (most deprived) | 1,293,838 | 1,005,179 (77.69%) | 31,990 (2.47%) | 861 (0.07%) | 255,808 (19.77%) |
| | 2 | 1,491,154 | 1,223,250 (82.03%) | 29,456 (1.98%) | 812 (0.05%) | 237,636 (15.94%) |
| | 3 | 1,665,132 | 1,446,137 (86.85%) | 25,368 (1.52%) | 903 (0.05%) | 192,724 (11.57%) |
| | 4 | 1,735,146 | 1,550,892 (89.38%) | 20,776 (1.20%) | 959 (0.06%) | 162,519 (9.37%) |
| | 5 (least deprived) | 1,815,919 | 1,656,249 (91.21%) | 16,177 (0.89%) | 1,183 (0.07%) | 142,310 (7.84%) |

***Figure S1. Cumulative percentage of patients recorded as declining a COVID-19 vaccination up to May 25th 2021, split by ethnicity, IMD, or age band.*** *Split by ethnicity for patients (a) aged 65+, (b) CEV/ At Risk, (c) aged 50-64; split by IMD for (d) aged 65+, (e) CEV/ At Risk, (f) aged 50-64; (g) split by age band for CEV / At Risk (CEV includes ages 16-69, At Risk includes ages 16-64).*

*a*

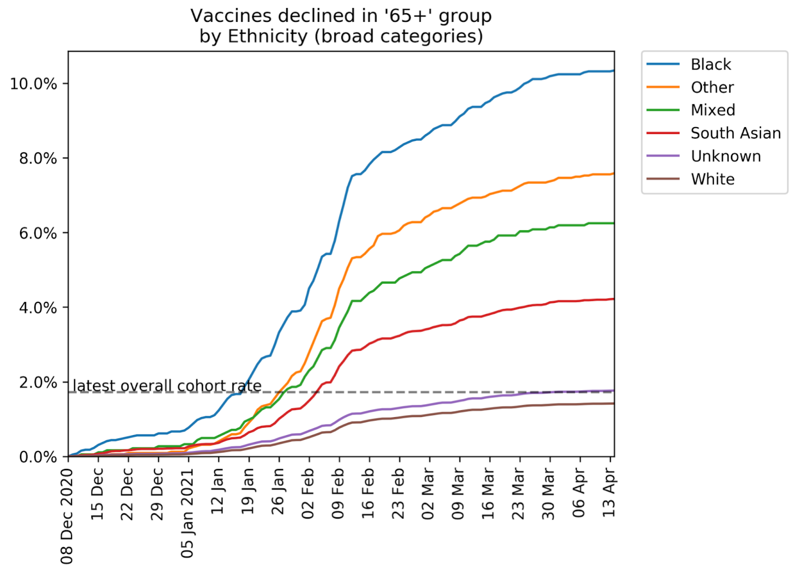


*b*

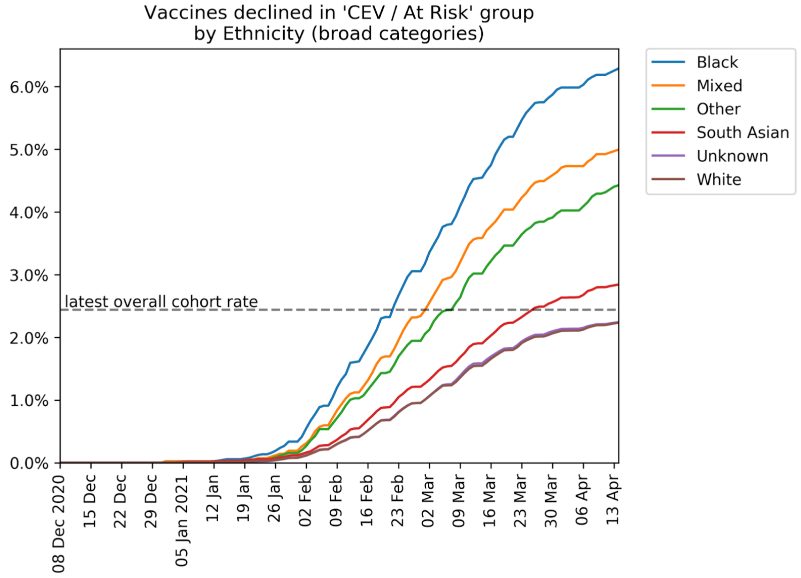

*c*

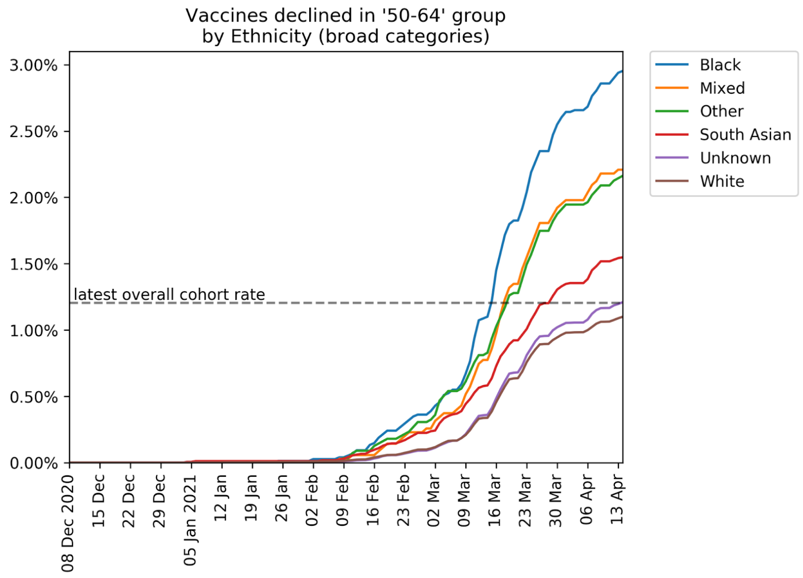

Vaccines declined in '50-64' group
by Ethnicity (broad categories)
3.00%
2.50%
2.00%
1.50%
1.00%
0.50%
0.00%
latest overall cohort rate
08 Dec 2020
15 Dec
22 Dec
29 Dec
05 Jan 2021
12 Jan
19 Jan
26 Jan
02 Feb
09 Feb
16 Feb
23 Feb
02 Mar
09 Mar
16 Mar
23 Mar
30 Mar
06 Apr
13 Apr
Black
Mixed
Other
South Asian
Unknown
White


*d*

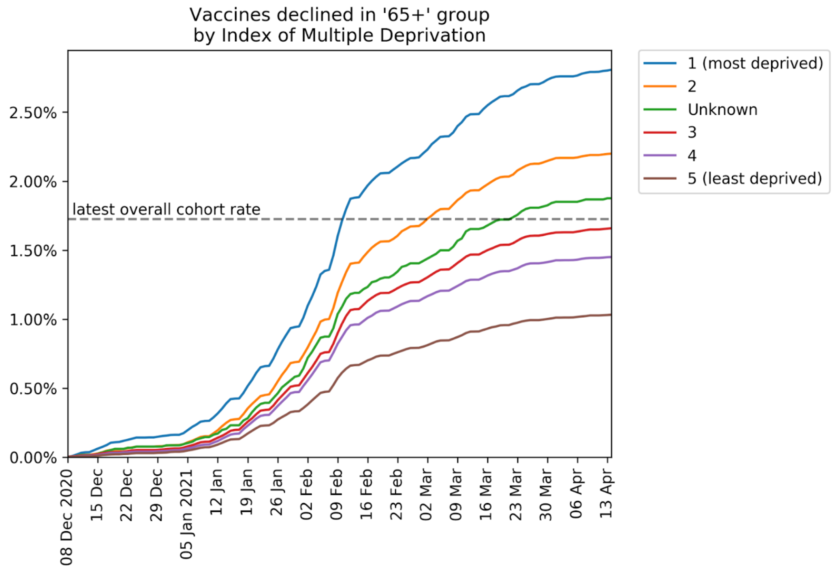

Vaccines declined in '65+' group
by Index of Multiple Deprivation
2.50%
2.00%
1.50%
1.00%
0.50%
0.00%
latest overall cohort rate
08 Dec 2020
15 Dec
22 Dec
29 Dec
05 Jan 2021
12 Jan
19 Jan
26 Jan
02 Feb
09 Feb
16 Feb
23 Feb
02 Mar
09 Mar
16 Mar
23 Mar
30 Mar
06 Apr
13 Apr
1 (most deprived)
2
Unknown
3
4
5 (least deprived)


*e*

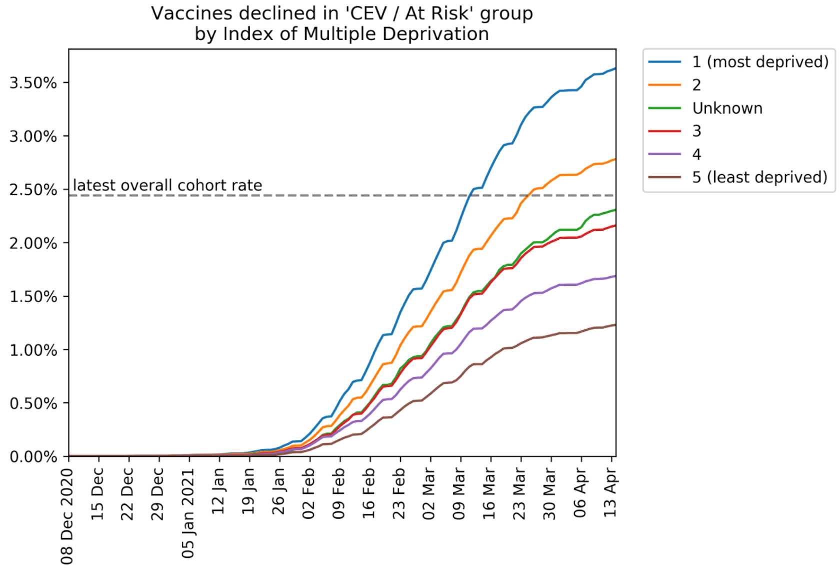

Vaccines declined in 'CEV / At Risk' group
by Index of Multiple Deprivation
3.50%
3.00%
2.50%
2.00%
1.50%
1.00%
0.50%
0.00%
latest overall cohort rate
08 Dec 2020
15 Dec
22 Dec
29 Dec
05 Jan 2021
12 Jan
19 Jan
26 Jan
02 Feb
09 Feb
16 Feb
23 Feb
02 Mar
09 Mar
16 Mar
23 Mar
30 Mar
06 Apr
13 Apr
1 (most deprived)
2
Unknown
3
4
5 (least deprived)

*f*

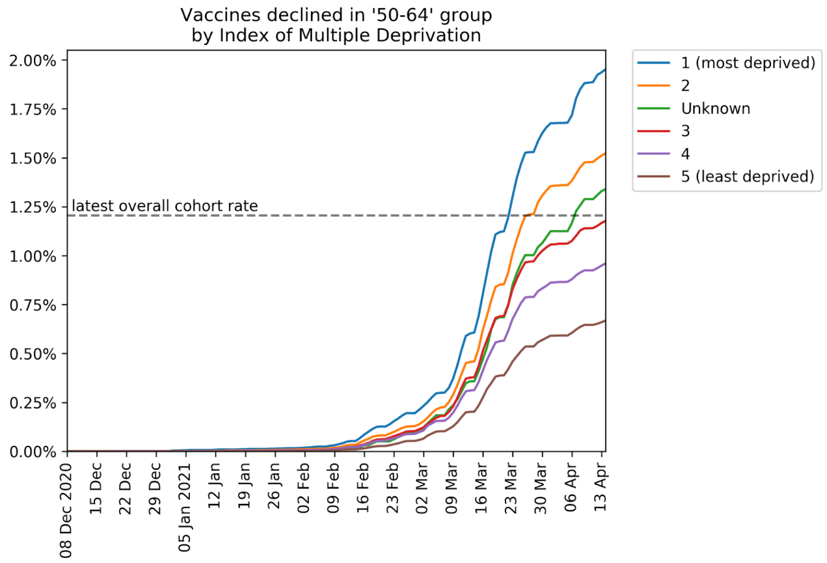
Vaccines declined in '50-64' group
by Index of Multiple Deprivation
latest overall cohort rate
1 (most deprived)
2
Unknown
3
4
5 (least deprived)
2.00%
1.75%
1.50%
1.25%
1.00%
0.75%
0.50%
0.25%
0.00%
08 Dec 2020
15 Dec
22 Dec
29 Dec
05 Jan 2021
12 Jan
19 Jan
26 Jan
02 Feb
09 Feb
16 Feb
23 Feb
02 Mar
09 Mar
16 Mar
23 Mar
30 Mar
06 Apr
13 Apr

*g*

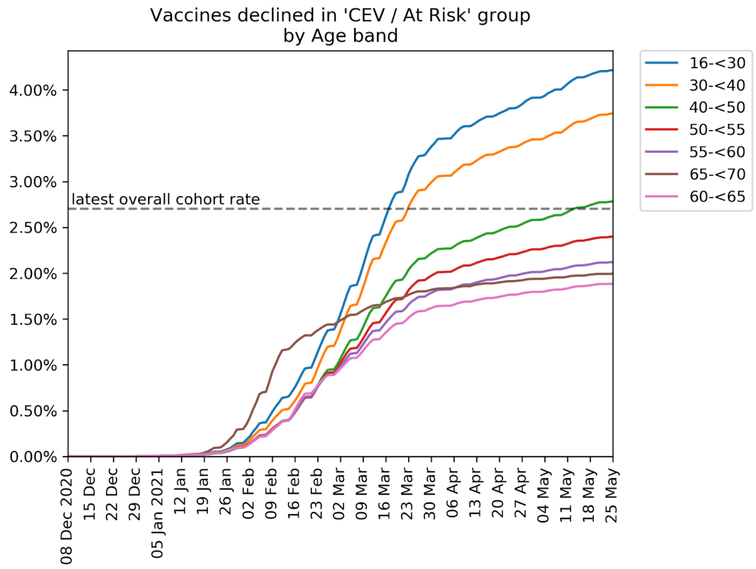
Vaccines declined in 'CEV / At Risk' group
by Age band
latest overall cohort rate
16-<30
30-<40
40-<50
50-<55
55-<60
65-<70
60-<65
4.00%
3.50%
3.00%
2.50%
2.00%
1.50%
1.00%
0.50%
0.00%
08 Dec 2020
15 Dec
22 Dec
29 Dec
05 Jan 2021
12 Jan
19 Jan
26 Jan
02 Feb
09 Feb
16 Feb
23 Feb
02 Mar
09 Mar
16 Mar
23 Mar
30 Mar
06 Apr
13 Apr
20 Apr
27 Apr
04 May
11 May
18 May
25 May

***Figure S2. Heatmap showing practice variation in the number of patients recorded as declining a COVID-19 vaccination per 1000 patients in priority groups, according to the total number of priority group patients in each practice, as at 25th May 2021.*** *Colour scale indicates number of practices (values of 4 may represent approximated counts). Non-linear axis scales are used: the largest category on the y-axis includes all values >100. Practices with 250 or fewer registered patients in priority groups and those with 10 or fewer vaccinated patients were excluded. Patients recorded as declined only includes unvaccinated patients.*

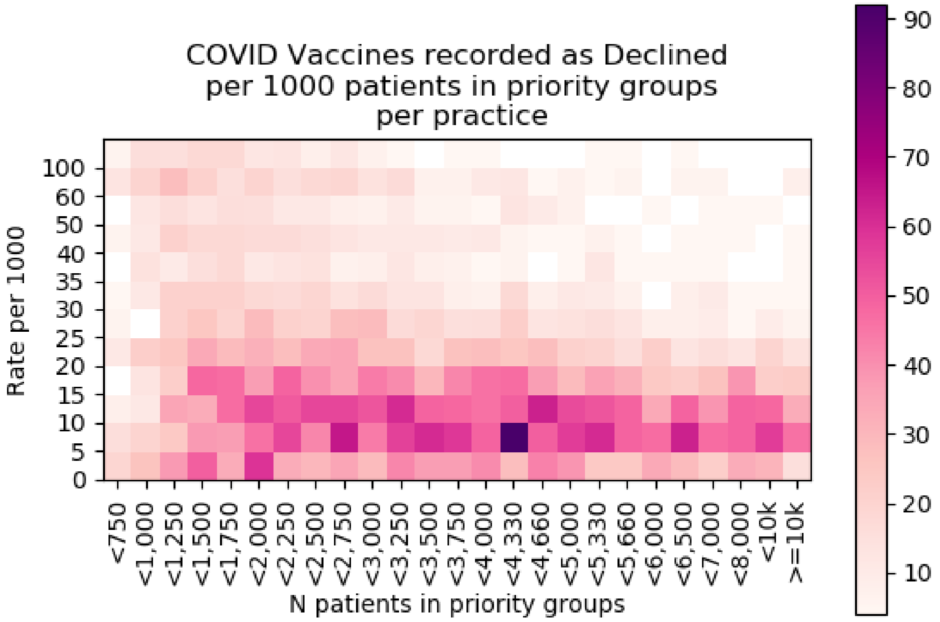
